# Response consistency of ChatGPT-4o for Type 2 Diabetes Nutrition and Physical-activity Recommendations: A Pilot NLP-based Assessment of GPT outputs

**DOI:** 10.64898/2026.06.23.26356399

**Authors:** Yundan Zhang, Xue-Jing Liu, Qiongzhi Hu, Karla I. Galaviz, Ines Gonzalez Casanova, Jason Colditz, Danny Valdez

## Abstract

Generative AI tools such as ChatGPT are increasingly used by the public to seek guidance on diet and physical activity for type 2 diabetes (T2D) prevention and management. However, the consistency of model outputs across different users and disease-stage scenarios remains insufficiently characterized. This pilot study aims to evaluate the word-level and semantic-level consistency of GPT-4o’s diet and physical activity responses for type 2 diabetes prevention and management. We designed 12 prompts covering four categories: prediabetes, diagnosed type 2 diabetes (T2D), diagnosed T2D with complications, and general questions that did not specify dysglycemia stage. Word-level similarity was quantified with Term Frequency-Inverse Document Frequency (TF-IDF) cosine scores; sentence-level semantic similarity was measured using large language models (LLMs) – DeBERTa-v3 MNLI to calculate the entailment probabilities. The results showed that mean cosine similarity across users was moderate (0.44–0.66), whereas mean entailment similarity was higher (0.68–0.81). Across stages, word-level similarity was low to moderate (0.28–0.63) and entailment similarity remained moderate to high (0.63–0.80). Low similarity commonly referenced distinct food choices, operational details, safety warnings, and stage-specific suggestions. GPT-4o generated semantically consistent but variably detailed responses and the moderate semantic variation suggested limited differentiation of response content across diabetes-related stages in this pilot consistency assessment.

**Author Summary:** This pilot study investigated the consistency of nutrition and physical activity recommendations generated by ChatGPT-4o for type 2 diabetes. While content accuracy is an important aspect of evaluating AI-generated health advice, answer consistency is also important, especially for medical-related guidance such as diabetes nutrition and lifestyle recommendations. We collected one-round responses from ChatGPT-4o and quantitatively compared the generated answers across users and prevention stages. Overall, ChatGPT-4o provided generally consistent recommendations on broad topics, including healthy eating, physical activity, weight management, and blood glucose monitoring. However, the operational details varied across users and stages, such as how recommendations were prioritized, framed, and translated into specific actions. More details were discussed in the paper. This study serves as a proof-of-concept showing that the consistency of AI-generated health recommendations can be measured quantitatively. Future work may expand this publicly available framework to follow-up conversations, larger sample sizes, more diverse user profiles, and further evaluation of accuracy and actionability.

## Introduction

Type 2 Diabetes (T2D) is a chronic condition impacting 38 million Americans as of 2021, with an additional 97.6 million experiencing prediabetes[1, 2]. When left unaddressed, prediabetes and T2D increase the likelihood for developing co-occurring and preventable conditions such as heart disease, vision impairment, and kidney disease[3, 4]. Therefore, proactive prevention and management are critical to slow disease progression and reduce the well-documented public health and medical burden of T2D in the US.

Lifestyle modification—such as improved diet and increased physical activity—are well-established strategies for preventing T2D, slowing T2D progression, and reducing T2D complications[5, 6]. However, many individuals face barriers to accessing conventional diabetes prevention and management care, including limited healthcare access in rural areas, high out-of-pocket costs, long wait times, and a shortage of culturally tailored resources[7–10]. These obstacles may deter timely preventive counseling and care, leading people to turn to freely accessible online communities, mobile apps— and increasingly Generative Artificial Intelligence (Gen-AI)— to research health information about T2D prevention and management[8, 11–13].

In 2025 as much as 60% of US adults used Gen-AI, a type of AI powered by Large Language Models (LLMs), in the last six months and 27% of adults reported using AI constantly[14]. These estimates may not reflect true use given that leading search engines including Google, Bing, and others now provide recommendations and suggestions that are powered by proprietary Gen-AI platforms. Yet, despite its wide accessibility and growing use, the reliability of Gen-AIs as a health coach and tool remains uncertain due to concerns about its effectiveness in providing consistent recommendations across users[15].

Indeed, research on human and Gen-AI interactions indicates that current Gen-AI limitations and human error can adversely impact responses. First, prior research shows that Gen-AI can confidently produce false or partially correct information, otherwise referred to as hallucinations[16, 17]. One study among people with food allergies found, for example, that ChatGPT - a leading Gen-AI platform commonly used for simple tasks - erroneously included allergens in 4 of 56 meal suggestions and failed to warn about energy-deficient plans, despite adhering to general dietary guidelines[18]. Similarly, another study found while ChatGPT could formulate an American Diabetes Association aligned diet for a T2D patient, advice was inconsistent and sometimes offered incorrect and conflicting advice upon repeated queries[19].

Additional investigations likewise suggest that human input can negatively impact the quality of information provided by Gen-AI, particularly if a user has no experience with a condition or activity. For example, AI-driven nutrition recommendations and physical activity guidance vary dramatically based on user input; and in many cases responses are inconsistent. These responses, in particular, may lack clinical specificity, omit critical details such as nutrient targets, or fail to provide safe recommendations for exercise duration[19, 20]. These well documented issues may, in turn, bias Gen-AI output or result in Gen-AI-provided recommendations that are not aligned with scientific standards or do not meet the current needs of an individual user.

Although AI has been applied to T2D management tasks such as meal planning and nutrition counseling[21, 22], only a small number of studies have specifically evaluated its ability to deliver *consistent* diet and physical activity guidance for people along the diabetes-care continuum, which includes prediabetes (i.e., prevention focus), T2D (i.e., management focus), and T2D with complications (i.e., advanced management with a tertiary prevention focus)[19, 20, 23, 24]. In other words, regardless of the degree of clinical accuracy of a single Gen-AI output, we do not know the extent to which multiple users are likely to encounter identical, similar, or dissimilar response patterns. Understanding the degree of alignment and divergence of responses is a critical first-step in understanding current gaps in Gen-AI utility for general use.

Existing research highlights notable concerns about current Gen-AI capabilities for such tasks, including inconsistent outputs across repeated queries, overly generalized recommendations, and partial incongruence with clinical guidelines[25–27]. However, these prior studies relied primarily on expert review of GPT outputs, with lack of quantitative or reproducible metrics to systematically assess the consistency of Gen-AI generated responses[19, 20, 24].

To address this gap, we pilot a new, natural language processing, machine learning, and LLM -supported framework to quantify the between user and across condition consistency of GPT-4o’s responses—which, at the time of writing was the most widely implemented GPT model— to dietary and physical activity questions posed by hypothetical Gen-AI users with T2D lifestyle questions. The specific objectives of this exploratory study were to:

1. Quantify across-user consistency by comparing word-level and semantic similarity between users;
2. Quantify stage-specific variation by analyzing word-level and semantic similarity across different T2D stages;
3. Identify and categorize sources of inconsistent stage-specific prevention and/or management strategies and provide future recommendations.

Through this formative assessment, we aim to identify the relative degree of alignment and divergence in content across a small number of hypothetical users. Findings from this study will inform the relative success of this framework in LLM response quantification and the degree of consistency across hypothetical users. Should the framework yield a reliable or sufficient signal, we expect to scale the results with larger sample sizes and hypothetical users—and move toward formal clinical assessment of responses in a bid to improve Gen-AI functionality for health information seeking.

## Results

### Descriptive Characteristics of the T2D Questionnaire

The raw sentence counts, means, and standard deviations for users (n=3) across six diet-related and six physical activity prompts are presented in **Table 1**. For diet-related prompts, the mean sentence counts ranged from 32-42 (SD = 3.00-6.81) for same prompt across users. Across prompts, the average number of sentences for each user ranged between 32-40 (SD = 3.93-5.05). For physical activity prompts, the average sentence counts ranged from 29-33 (SD = 2.08-8.39) across users, while within-user sentence counts averaged 25-33 (SD = 2.94-5.96).

**Table 1.**
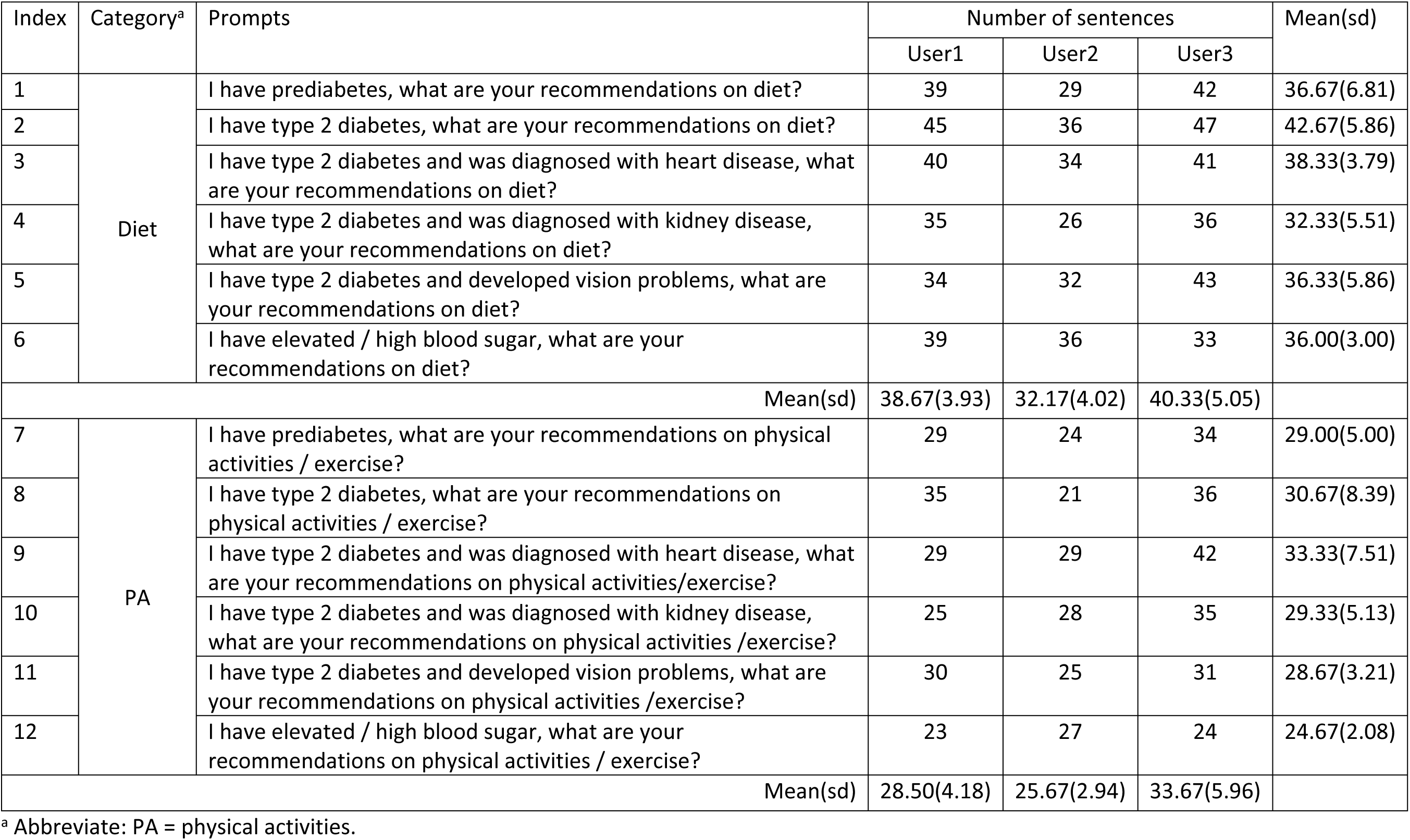
Prompt design and categories, sentence counts, and descriptive statistics.

| Index | Category<sup>a</sup> | Prompts | Number of sentences |  |  | Mean(sd) |
| --- | --- | --- | --- | --- | --- | --- |
|  |  |  | User1 | User2 | User3 |  |
| 1 | Diet | I have prediabetes, what are your recommendations on diet? | 39 | 29 | 42 | 36.67(6.81) |
| 2 |  | I have type 2 diabetes, what are your recommendations on diet? | 45 | 36 | 47 | 42.67(5.86) |
| 3 |  | I have type 2 diabetes and was diagnosed with heart disease, what are your recommendations on diet? | 40 | 34 | 41 | 38.33(3.79) |
| 4 |  | I have type 2 diabetes and was diagnosed with kidney disease, what are your recommendations on diet? | 35 | 26 | 36 | 32.33(5.51) |
| 5 |  | I have type 2 diabetes and developed vision problems, what are your recommendations on diet? | 34 | 32 | 43 | 36.33(5.86) |
| 6 |  | I have elevated / high blood sugar, what are your recommendations on diet? | 39 | 36 | 33 | 36.00(3.00) |
| Mean(sd) |  |  | 38.67(3.93) | 32.17(4.02) | 40.33(5.05) |  |
| 7 | PA | I have prediabetes, what are your recommendations on physical activities / exercise? | 29 | 24 | 34 | 29.00(5.00) |
| 8 |  | I have type 2 diabetes, what are your recommendations on physical activities / exercise? | 35 | 21 | 36 | 30.67(8.39) |
| 9 |  | I have type 2 diabetes and was diagnosed with heart disease, what are your recommendations on physical activities/exercise? | 29 | 29 | 42 | 33.33(7.51) |
| 10 |  | I have type 2 diabetes and was diagnosed with kidney disease, what are your recommendations on physical activities /exercise? | 25 | 28 | 35 | 29.33(5.13) |
| 11 |  | I have type 2 diabetes and developed vision problems, what are your recommendations on physical activities /exercise? | 30 | 25 | 31 | 28.67(3.21) |
| 12 |  | I have elevated / high blood sugar, what are your recommendations on physical activities / exercise? | 23 | 27 | 24 | 24.67(2.08) |
| Mean(sd) |  |  | 28.50(4.18) | 25.67(2.94) | 33.67(5.96) |  |

<sup>a</sup> Abbreviate: PA = physical activities.

### Across-user word-level and semantic-level consistency

We observed low to moderate word level and moderate to high semantic level similarity across all user pairwise comparisons. **Table 2** presents scores grouped by diet and physical activity categories. For diet related prompts (Indexes 1-6), word-level similarity across pairwise comparisons was moderate (overall range 0.44–0.66), with per-prompt means across users ranging from as low as 0.49 (Index 1) to as high as 0.66 (Index 4). Semantic-level similarity scores for diet-related prompts were moderate to high (overall range 0.68–0.79), with per-prompt means across users ranging from as low as 0.68 (Index 5) to as high as 0.79 (Index 3). PA prompts (Indexes 7–12) were similarly mixed, with across-users word-level means ranging from 0.44 (Index 11) to 0.60 (Index 10). Likewise semantic level means for physical activity prompts across users were higher and ranged from 0.72 (Index 8) to 0.81 (Index 11).

**Table 2.** Word-level and semantic similarity across users. Word-level similarity was low-to-moderate across all comparisons; semantic similarity was moderate-to-high across all comparisons. This suggests users receive similar content phrased differently.

| Index | Category<sup>a</sup> | Word-level similarity |  |  |  | Semantic-level similarity |  |  |  |
| --- | --- | --- | --- | --- | --- | --- | --- | --- | --- |
|  |  | User1/User2 | User1/User3 | User2/User3 | Mean(sd) | User1/User2 | User1/User3 | User2/User3 | Mean(sd) |
| 1 | Diet | 0.54 | 0.50 | 0.42 | 0.49(0.06) | 0.72 | 0.77 | 0.73 | 0.74(0.03) |
| 2 |  | 0.55 | 0.51 | 0.52 | 0.53(0.02) | 0.72 | 0.79 | 0.72 | 0.74(0.04) |
| 3 |  | 0.60 | 0.56 | 0.58 | 0.58(0.02) | 0.78 | 0.84 | 0.75 | 0.79(0.05) |
| 4 |  | 0.61 | 0.70 | 0.68 | 0.66(0.05) | 0.73 | 0.84 | 0.72 | 0.76(0.07) |
| 5 |  | 0.61 | 0.59 | 0.60 | 0.60(0.01) | 0.69 | 0.69 | 0.66 | 0.68(0.02) |
| 6 |  | 0.48 | 0.52 | 0.48 | 0.50(0.02) | 0.74 | 0.75 | 0.72 | 0.74(0.02) |
|  | Mean(sd) | 0.56 (0.05) | 0.56 (0.08) | 0.54 (0.09) |  | 0.73 (0.03) | 0.78 (0.06) | 0.72 (0.03) |  |
| 7 | PA | 0.55 | 0.52 | 0.57 | 0.55(0.03) | 0.82 | 0.81 | 0.71 | 0.78(0.06) |
| 8 |  | 0.57 | 0.59 | 0.49 | 0.55(0.05) | 0.67 | 0.85 | 0.65 | 0.72(0.11) |
| 9 |  | 0.56 | 0.51 | 0.54 | 0.54(0.03) | 0.86 | 0.72 | 0.74 | 0.77(0.08) |
| 10 |  | 0.59 | 0.55 | 0.65 | 0.60(0.05) | 0.84 | 0.74 | 0.79 | 0.79(0.05) |
| 11 |  | 0.47 | 0.43 | 0.41 | 0.44(0.03) | 0.82 | 0.85 | 0.76 | 0.81(0.05) |
| 12 |  | 0.55 | 0.51 | 0.48 | 0.51(0.04) | 0.78 | 0.80 | 0.73 | 0.77(0.04) |
|  | Mean(sd) | 0.55 (0.04) | 0.52 (0.05) | 0.52 (0.08) |  | 0.80 (0.07) | 0.79 (0.05) | 0.73 (0.05) |  |

<sup>a</sup> Abbreviate: PA = physical activities.

Our findings are further supported by **Figure 1 and S3**, which visualize findings in sentence level similarity. As shown in **S3**, Index 5 (T2D+DR) shows a lower mean similarity score across users compared to Index 1 (Pre-D, 0.09 [0.02, 0.13]), Index 2 (T2D, 0.06 [0.02, 0.10]), Index 3 (T2D+CVD, 0.08 [0.05, 0.14]), and Index 4 (T2D+CKD, 0.10 [0.06, 0.15]). In addition, the general stage-level prompts for diet (Index 6) and physical activity (Index 12) showed lower mean across-user similarity than the more specific prompts, including Index 3 and Index 4, as well as Index 7 through Index 11. **Figure 1** also shows a slightly higher overall similarity score and lower variance for physical activity recommendation compared to diet recommendations. Distribution of low, median, and high paired similarity are reported in **S1**, while paired sentence examples across threshold are displayed in **Supplementary table 6 S6**.

**Figure 1.**
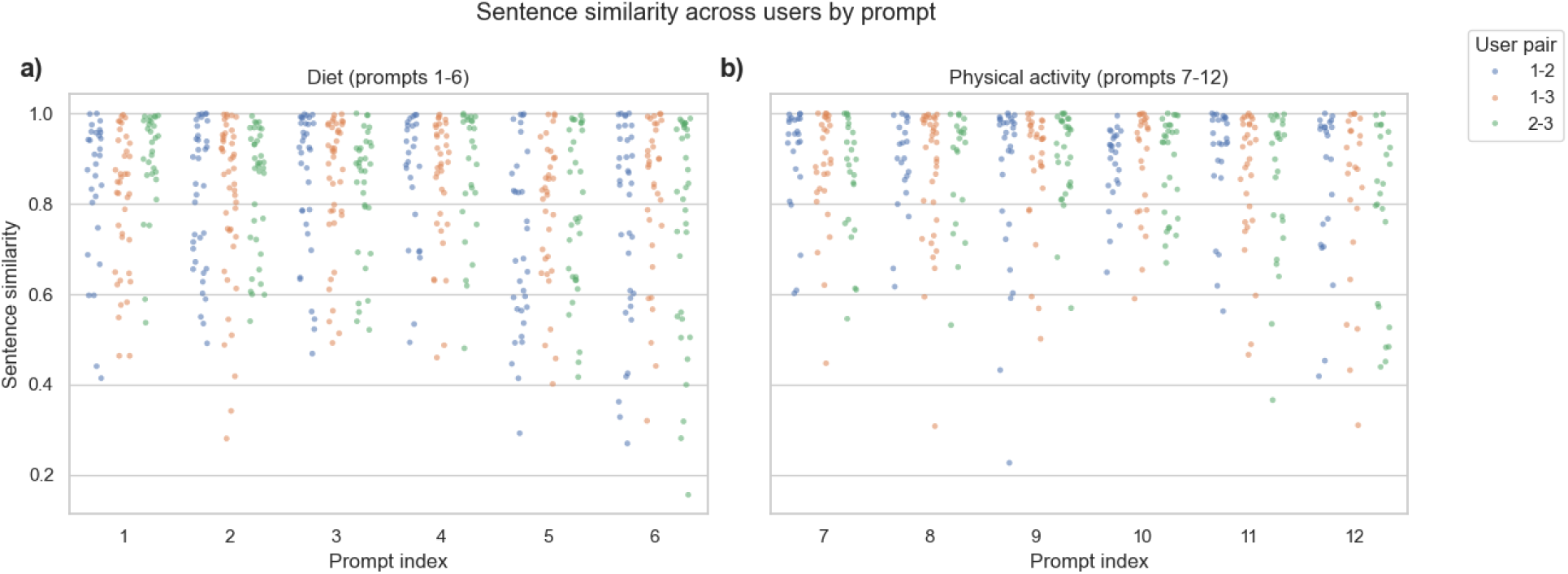
Sentence-level semantic similarity across users by prompt^1^ ^1^ Strip plots of similarity score for each of the 12 prompts. a) Similarity score across three users for diet prompts (Index 1-6). b) Similarity score across three users for physical activity prompts (Index 7-12). Higher scores denote greater sentence-level agreement across the three users.

We also observed low or weakly matched sentence pairs across users, which are provided in **S5**. For comparisons pertaining to diet, sentences with weak or unmatched similarity revealed several common divergences, including: (1) lists, menus, and itemized food guidance (such as sample meals, specific breakfast/lunch/dinner/snack, and named food such as berries, salmon, quinoa, etc.); (2) condition-specific constraints (kidney or vision comorbidities rules such as attention to potassium, antioxidants, lutein, etc.); (3) operational details (supplement intake such as recommendation for omega-3, magnesium, cinnamon, etc., food label-checking, and routine monitoring); (4) mechanistic explanations (glycemic spikes, insulin response, and alcohol interactions); and (5) safety and referral warnings.

Physical activity prompts show the same pattern but with fewer mismatches. Weakly matched or unmatched sentences emphasized: (1) specific activities (e.g., yoga, Tai Chi); (2) operational details (time of activity such as “3-5 minutes after each meal”, session structuring, and use of adaptive equipment); (3) safety and clinical guidance (concrete safety note such as “cool-down physiology”, consulting with providers, blood pressure or glucose monitoring, foot care routines, etc.); (4) condition-specific guidance (kidney or vision comorbidities safety constraints such as “avoid heavy lifting”, “monitor ketone generation”, etc.). Mid-similarity pairs convey guideline-consistent advice in different units or phrasings (minutes vs. qualitative terms like “short warm-up” or “light walk”) and vary in methods for gauging physical activity intensity (heart rate targets vs. the talk test). High-similarity sentences cover regular moderate-intensity aerobic activity, often with added resistance training, alongside consistent glucose monitoring.

### Across disease-stage word-level and semantic-level consistency

**Table 3** highlights word and semantic similarity across T2D stages grouped by diet and physical activity. In alignment with our findings across users, the word-level similarities for diet prompts across T2D stages are low to moderate, with means for each stage pair spanning 0.28–0.63. The lowest similarity was observed in the T2D+CKD and T2D+DR comparison, while highest similarity in the Pre-D and T2D comparison. Semantic-level similarities for diet prompts were consistently moderate to high, with means across users spanning 0.63–0.80. The lowest was found in the T2D+CKD and T2D+DR comparison, while highest was found in Pre-D and T2D comparison. The similarity for physical activity across stages is consistently higher than diet recommendations. For physical activity prompts, word-level means across users ranged from 0.33–0.55 (lowest for the Pre-D and T2D+CKD, T2D+CKD and T2D+DR comparisons; and highest for Pre-D/T2D comparisons), and semantic means were notably and consistently high across stages ranging from 0.74–0.80 (lowest for the T2D, T2D+CKD and T2D, T2D+DR comparison; highest for the Pre-D and T2D comparison).

**Table 3.** Word-level and semantic similarity across T2D progression stages. Word-level similarity was low to moderate, with notable declines in word-level similarity in diet prompts. Semantic similarity was moderate to high, with notable increases in similarity in physical activity prompts. While we observed a decline in similarity in comparisons by prevention-stage and management-stage T2D, these declines were less than expected and may have hinged on operational details.

| Category<sup>a</sup> | Status pair<sup>b</sup> | Word-level similarity |  |  |  | Semantic-level similarity |  |  |  |
| --- | --- | --- | --- | --- | --- | --- | --- | --- | --- |
|  |  | User1 | User2 | User3 | Mean (sd) | User1 | User2 | User3 | Mean (sd) |
| Diet | Pre-D/T2D | 0.61 | 0.70 | 0.57 | 0.63(0.07) | 0.73 | 0.74 | 0.80 | 0.76(0.04) |
|  | Pre-D/T2D+CVD | 0.61 | 0.56 | 0.58 | 0.58(0.03) | 0.77 | 0.73 | 0.85 | 0.78(0.06) |
|  | Pre-D/T2D+CKD | 0.36 | 0.35 | 0.31 | 0.34(0.03) | 0.68 | 0.67 | 0.73 | 0.69(0.03) |
|  | Pre-D/T2D+DR | 0.47 | 0.35 | 0.51 | 0.44(0.08) | 0.64 | 0.66 | 0.79 | 0.69(0.08) |
|  | T2D/T2D+CVD | 0.56 | 0.61 | 0.54 | 0.57(0.04) | 0.74 | 0.71 | 0.79 | 0.75(0.04) |
|  | T2D/T2D+CKD | 0.36 | 0.37 | 0.28 | 0.34(0.04) | 0.69 | 0.58 | 0.65 | 0.64(0.06) |
|  | T2D/T2D+DR | 0.41 | 0.47 | 0.51 | 0.46(0.05) | 0.63 | 0.66 | 0.71 | 0.67(0.04) |
|  | T2D+CVD/T2D+CKD | 0.42 | 0.43 | 0.36 | 0.40(0.04) | 0.75 | 0.66 | 0.72 | 0.71(0.05) |
|  | T2D+CVD/T2D+DR | 0.49 | 0.47 | 0.46 | 0.48(0.01) | 0.68 | 0.64 | 0.72 | 0.68(0.04) |
|  | T2D+CKD/T2D+DR | 0.31 | 0.31 | 0.23 | 0.28(0.05) | 0.67 | 0.59 | 0.64 | 0.63(0.04) |
|  | Mean(sd) | 0.46(0.11) | 0.46(0.13) | 0.44(0.13) |  | 0.70(0.05) | 0.66(0.05) | 0.74(0.07) |  |
| PA |  |  |  |  |  |  |  |  |  |
|  | Pre-D/T2D | 0.60 | 0.49 | 0.57 | 0.55(0.05) | 0.82 | 0.75 | 0.84 | 0.80(0.05) |
|  | Pre-D/T2D+CVD | 0.45 | 0.45 | 0.52 | 0.47(0.04) | 0.81 | 0.75 | 0.78 | 0.78(0.03) |
|  | Pre-D/T2D+CKD | 0.36 | 0.32 | 0.33 | 0.34(0.02) | 0.77 | 0.73 | 0.75 | 0.75(0.02) |
|  | Pre-D/T2D+DR | 0.41 | 0.27 | 0.41 | 0.36(0.08) | 0.83 | 0.68 | 0.76 | 0.76(0.08) |
|  | T2D/T2D+CVD | 0.51 | 0.44 | 0.55 | 0.50(0.05) | 0.76 | 0.71 | 0.81 | 0.76(0.05) |
|  | T2D/T2D+CKD | 0.38 | 0.36 | 0.43 | 0.39(0.03) | 0.70 | 0.73 | 0.78 | 0.74(0.04) |
|  | T2D/T2D+DR | 0.42 | 0.37 | 0.53 | 0.44(0.08) | 0.79 | 0.68 | 0.76 | 0.74(0.06) |
|  | T2D+CVD/T2D+CKD | 0.37 | 0.39 | 0.39 | 0.38(0.01) | 0.79 | 0.81 | 0.77 | 0.79(0.02) |
|  | T2D+CVD/T2D+DR | 0.41 | 0.29 | 0.43 | 0.38(0.07) | 0.80 | 0.73 | 0.73 | 0.75(0.04) |
|  | T2D+CKD/T2D+DR | 0.30 | 0.30 | 0.39 | 0.33(0.05) | 0.76 | 0.72 | 0.80 | 0.76(0.04) |
|  | Mean(sd) | 0.42(0.08) | 0.37(0.07) | 0.46(0.08) |  | 0.78(0.04) | 0.73(0.04) | 0.78(0.03) |  |

<sup>a</sup> Abbreviate: PA = physical activities.
<sup>b</sup> “A/B” denotes pairwise similarity between condition A and condition B. Abbreviates: Pre-D = prediabetes; T2D = type 2 diabetes; CVD = cardiovascular disease; CKD = chronic kidney disease; DR = diabetic retinopathy.

Our findings are further supported by **Figure 2 and S4**, which shows the distribution of sentence level similarity across the stage. Responses similarity between pre-diabetes and T2D was higher than similarity between T2D with complications. A higher similarity score is also found between Pre-D and T2D+CVD compared to similarity between later T2D stages (T2D+CVD, T2D+CKD, and T2D+DR) as shown in **Figure 2 and S4.** Distribution of low, median, and high sentence similarity across stages are reported in **S2,** while paired sentence examples across threshold are displayed in **S6**.

**Figure 2.**
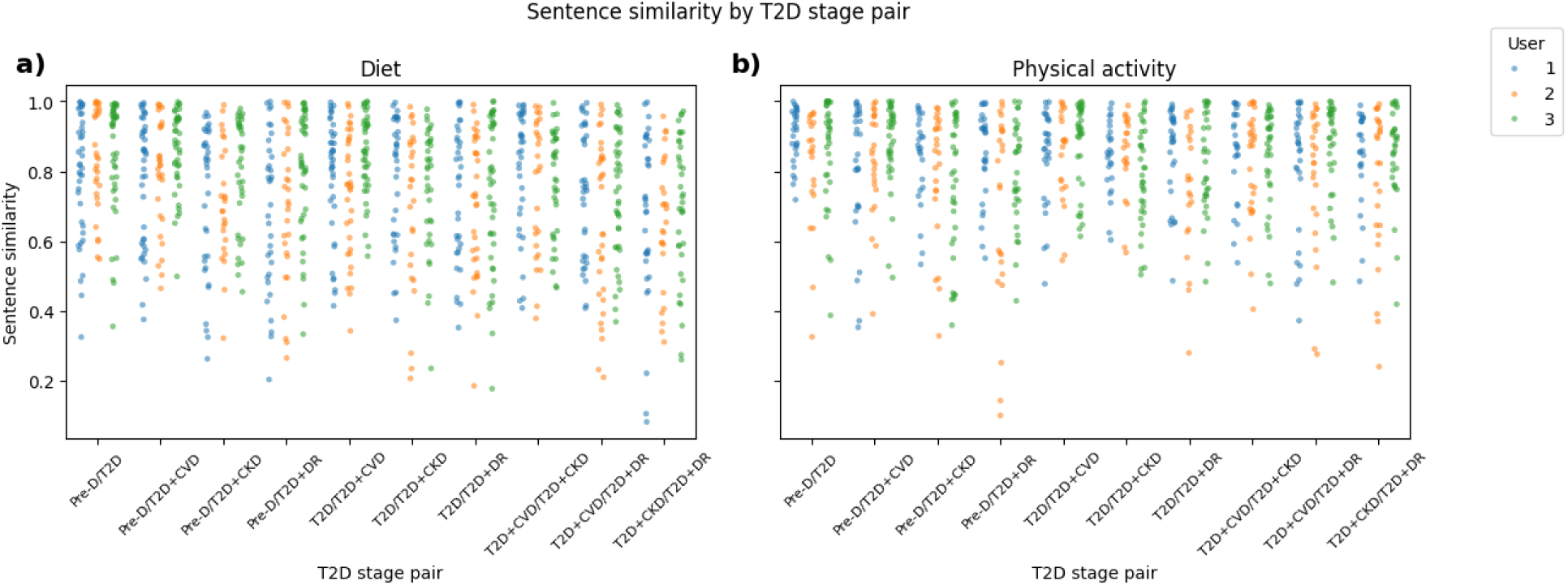
Sentence-level semantic similarity across paired T2D stage^1, 2^ ^1^ Strip plots of similarity score across the paired T2D stages aggregated from three users. a) Similarity score between paired stages for dietary prompts. The plot shows similarity score from GPT-4o responses various across different stages pairs. b) Similarity score between paired stages for physical activity prompts. ^2^ Abbreviations: Pre-D = prediabetes; T2D = type 2 diabetes; CVD = cardiovascular disease; CKD = chronic kidney disease; DR = diabetic retinopathy.

Across stages, dietary and physical activity recommendations showed the same weak/unmatched patterns seen in the across-user analysis (**S5**). Weak and unmatched sentence pairs were concentrated in condition-specific content and operational details. Stage-related differences are most pronounced in T2D+DR and T2D+CKD prompts but less evident for T2D+CVD, where advice and similarity scores aligned more closely with general Pre-D and T2D responses.

For CVD, semantic agreement across users was high—nearly all responses advised limiting sodium—yet the operational details varied: some warned broadly about processed foods, others recommended cooking at home, substituting spices for salt, or avoiding frozen meals. For kidney disease, answers were more explicitly condition-specific (protein management, portion control, sodium limits, and phosphorus/potassium restrictions) but differed in execution. Some recommendations emphasized lowering total protein and preferring plant-based sources; others prioritized “adequate protein” with sample plate proportions; some explained the rationale for protein restriction and advised clinician consultation in later kidney disease stage. For vision complications, we found the greatest divergence across users and the highest number of condition-specific suggestions. Recommendations mentioned vitamins A/E/C to varying degrees, antioxidants with or without explicit lutein/zeaxanthin guidance, omega-3s from food versus fish-oil supplements, occasional mentions of magnesium, and inconsistent advice on eye-exams and referrals.

For physical activity prompts involving T2D with complications, responses more frequently included medical clearance and monitoring language, especially for kidney and vision disease—with kidney-specific cautions (e.g., avoiding heavy lifting, managing fluids) and vision-adapted supports (e.g., stationary or anchored modalities, audio cues, an exercise partner).

## Discussion

A growing proportion of US adults now see more value in receiving health advice directly from Gen-AI models, citing convenience, physician mistrust, and historic inequity in healthcare access[28–31]. We tested an advanced machine learning pipeline capable of measuring GPT-4o’s ability to provide consistent recommendations for T2D prevention and management.

Overall, we observed that GPT-4o produced moderate word similarity and moderate-to-high semantic similarity across all pairwise comparisons. These results suggest that our word-level and semantic results diverged in anticipated ways. TF-IDF (or word) similarity is sensitive to paraphrasing, list ordering, and response length. Therefore, responses that convey similar meaning using different phrasing or formatting may receive lower word-level similarity scores. In contrast, entailment can remain high even when sentence structure differs. This indicates that responses often preserved overlapping high-level meaning despite differences in wording and structure.

However, despite these anticipated differences stemming from study design, we observed variations in T2D user-specific and stage-specific operational details. Because responses were generated through the public ChatGPT interface, where generation parameters could not be controlled, these variations reflect a user-facing interface variability. This, in turn, suggests output through ChatGPT interface may not necessarily imply consistent actionability. This pilot assessment cannot be interpreted as a controlled evaluation of model behavior under fixed generation settings. We contextualize our findings further, below.

### Contextualizing Consistent and Inconsistent T2D Dietary and Physical Activity Recommendations

Highly similar sentences often reflect broad diabetes statements (e.g., “managing diabetes helps control blood sugar”) and generic recommendations where there is strong agreement on a balanced, low-glycemic index, high-fiber dietary pattern with healthy fats and lean proteins. Common recommendations observed for physical activities included engaging in moderate aerobic and strength building physical activities, and to consult with physicians before making major lifestyle changes.

Conversely, we identified several inconsistencies in GPT-4o’s response schema. While moderate to high similarity across users may equate to generally consistent responses, specifics varied across users, particularly in lists/menus, condition-specific constraints, operational details, mechanisms, and safety/referral notes. These mismatches suggest that within the same prompt, responses can vary in detail and justification. For example, a query about T2D with kidney disease may yield one answer emphasizing supplements to reduce renal burden while another offers a different strategy such as consuming dark green vegetables.

Paired sentences with median level similarity tend to share general healthy eating themes (portion control, hydration, fiber emphasis, avoiding processed foods, etc.) and behavioral tips (“use a smaller plate”, “limit alcohol”, “monitor blood sugar”, and others.), differing mainly in phrasing or depth. In some cases, details diverged—meaning examples were different and mutually exclusive by user. Yet, in others, these details were omitted entirely, resulting in overly general answers. These inconsistencies, although subtle and not unexpected for general purpose LLMs such as GPT-4o, may illustrate documented limitations in delivering consistent guidance, notably among key details that may directly influence user behavior and decision making. This aligns with broader cautions about using LLMs in health contexts without safeguards[32].

It is also noteworthy that responses showed limited differentiation by disease-progression stage. Although national guidelines do not differ substantially between prediabetes and diagnosed T2D, advanced T2D with complications requires more person-specific adaptation beyond general recommendations to prevent tertiary comorbidities[33]. In comparisons between prediabetes and T2D with complications responses, we observed only a modest decline in similarity levels. This decline may reflect some variation in response content related to disease progression; however, the remaining high contextual overlap suggests that responses may not have been strongly differentiated across prevention and later-stage management contexts. These findings raise questions about the extent to which general-purpose LLM outputs adapt recommendations to more complex T2D scenarios.

### Study Implications and Future Directions

Our findings support that observed response consistency alone on a time-stamped snapshot is insufficient to establish clinical utility. Because this study does not evaluate accuracy against clinical guidelines or expert judgment, we cannot conclude whether the recommendations are clinically appropriate. Instead, we interpret the results as evidence that GPT-4o can generate semantically consistent responses while varying in the operational details that users may rely on for behavior modification. These issues may be particularly salient for individuals with complex T2D presentations or late-stage complications, where missing context and variable specificity could lead to misinterpretation.

Future work in this area should consider scaling this evaluation by increasing the number of users for comparison, incorporating clinically richer hypothetical profiles, and extending the design to multi-turn follow-up interactions. Where feasible, future studies should also replicate the analysis using API-based access with fixed generation parameters and repeat evaluations at multiple time points to quantify temporal drift as models are updated. For example, hypothetical users may begin with simple queries and slowly add details that were not included in this work, such as blood work, test results, known adaptations, weight, BMI, and biomarker profiles[34]. Recording each response and measuring consistency with the pipeline used in this study could determine whether these details lead to sufficient divergence in responses.

While several recent studies evaluating Gen-AI in clinical contexts highlight limitations in delivering high-quality nutrition and physical-activity guidance, they also underscore that guideline concordance and clinical safety are distinct evaluation dimensions from response consistency[35, 36]. Accordingly, we recommend additional content analytic work for clinical safety, such as including diabetes and nutrition expert to rate the quality of responses along clinical lines (e.g. guideline concordance, safety critical content, potential harmful or misleading information, and completeness and actionability) along clinical guidelines from major nutrition authorities, including the American Dietetic Association and others.

Finally, we also propose others replicate our study across different AI models, including Claude, Grok, and Gemini, and with larger sample sizes. Ideally these studies continue to utilize public-facing chat bot interfaces to simulate actual real-world use.

### Limitations

Our study is subject to limitations. First, we queried GPT-4o through the public interface rather than the API to reflect typical end-user conditions for ecological validity. However, the interface does not allow control or reporting of generation parameters, model-routing settings, system prompts, or other system-level settings, so some variability may reflect stochastic sampling rather than stable model behavior. Second, our study used single-turn, general lay-language prompts, which may not reflect the quality of multi-turn follow-up interactions or the degree of tailoring that could occur when users provide more detailed personal context. Given the population health literacy levels of only 12%, we intentionally used general queries to simulate realistic health-seeking behavior in Gen-AI tools and AI-enabled search engines[37]. At the same time, lay queries may lack the clinical specificity required for a model to generate highly individualized and actionable guidance[38]. Third, relying solely on sentence-level entailment can overlook broader contextual content within paragraphs. The variability might be affected by the length of sentences and segmentation methods we applied. However, as stated in multiple entailment similarity studies, the assessment for paragraph level similarity remains challenging especially for long content[39, 40]. Grouping sentences by thematic units or calibrating entailment thresholds against human similarity judgments might yield a more complete picture of response consistency, while the methods for noisy control and accuracy is still under discussion. Fourth, our number of users was intentionally small because the public-interface data collection process was manual and the DeBERTa sentence-pair workflow required substantial pairwise inference. Therefore, generalizability to real-world users is limited and findings should be interpreted as proof-of-concept evidence. Fifth, our study didn’t address the credibility for clinical alignment, and the interpretation of higher similarity is not considered as clinical appropriateness or safety.

### Conclusions

This study provides a pilot, quantitative assessment of GPT-4o’s response consistency for T2D lifestyle recommendations. Our findings reinforce the need for additional safeguards that more effectively control for GenAI health seeking and response variability[41]. Such safeguards can range from additional review of content or through additional computer engineering to adapting models for more refined tasks. Potentially useful approaches include fine-tuned LLMs with controlled reasoning trained on T2D clinical recommendations, user health data, and outcome-linked ground truth[42, 43]; schema-based structured outputs[44]; a user-guidance intake layer that gathers essential context (stage, labs, medications, functional limits, preferences)[45]; and retrieval-augmented generation (RAG) to ground recommendations in scientific sources[46]. These approaches would ensure models are equipped to handle diverse user needs and deliver stage-specific and scientifically credible recommendations. By moving toward a sustainable framework for effective GenAI health seeking, and improving response quality, clinicians may be more equipped to consider GenAI’s role in 21st century medical care.

## Methods

### Prompt Design

We designed a 12-item prompt set with input from diabetes and nutrition experts using lay language. The prompts were designed to simulate actual queries related to dietary and physical activity questions that people with T2D may pose in Gen-AI or Internet search engines with AI features, such as Google. The prompts were designed to reflect T2D prevention and management: prevention-focused prediabetic (Pre-D), management-focused T2D, and tertiary prevention-focused T2D with complications, which includes T2D with end-organ or vascular outcomes. For the late-stage T2D with complications category, we specifically included three common T2D-related comorbidities: cardiovascular disease (T2D+CVD), chronic kidney disease (T2D+CKD), and diabetic retinopathy (T2D+DR)[47]. Additionally, a fourth prompt category was created to represent general scenarios that did not specify any dysglycemia stage or complication. The full 12-prompt inventory is available in **Table 1**. All prompts were submitted to ChatGPT interface exactly as shown in **Table 1**.

### Data Collection

We collected our data using the ChatGPT-4o model, developed by OpenAI (Version: *gpt-4o-2024-08-06,* November 2024). To simulate user interactions, three research assistants independently submitted the same prompts using separate, anonymous ChatGPT accounts and temporary chat windows. Prompts were submitted sequentially during the same data-collection window and from the same geographic region. Each prompt was entered in a new, temporary chat session to minimize the contextual memory effects from previous interactions that tailored to each individual. No additional interface functions, such as file upload, image input, or web-browsing tools, were intentionally used. As a formative study, sample size was intentionally small due to computational processing constraints.

### Data preprocessing

We conducted two levels of analysis – word-level and semantic level. Word-level similarity measures the exact overlap of words and phrases, while the semantic similarity measures the contextual overlap, even if the words and phrasing are different (i.e., replying with different words that ultimately convey the same meaning).

The data were preprocessed separately for each of the two levels of analysis, using Python (v3.12) with NLTK(v3.9.3) for text cleaning. For the word-level similarity analysis, we processed each generated answer using the following steps: (1) convert all words into lowercase, (2) remove punctuation and numbers, and (3) remove stop words and lemmatize. For our semantic similarity analysis, each response was segmented into clean sentences. We first tokenized each raw, unedited answer with *sent_tokenize*, then further split on line breaks and removed any fragments that consisted of numbers, symbols, or whitespace. Short segments (fewer than four words) were excluded.

### Data Analysis

The data analyses for both word-level and semantic-level similarity were conducted across two dimensions: responses from different users given the same prompt (across-user similarity) and responses from the same user across different disease stages (across-stage similarity). All analyses were performed in Python (v3.12). We used scikit-learn (v1.7.0) for TF-IDF vectorization and cosine similarity; Hugging Face transformers (v4.53.2) with PyTorch (v2.7.1) to run a DeBERTa-v3 MNLI entailment model (potsawee/deberta-v3-large-mnli) for semantic similarity.

### Word Level Similarity Analysis

We used TF-IDF score to embed the text to calculate the word-level similarity. The TF-IDF score is the product of a term frequency (TF) score and inverse document frequency (IDF) score. TF measures how frequently a term occurs in a document, while IDF measures the importance of a term by weighting the same term’s frequency across all other documents[48]. Responses generated by GPT-4o were embedded into vector representations using TF-IDF, and cosine similarity was applied to measure the distance between vectors as an indicator of word-level similarity[49]. For across-user similarity, the corpus was defined as all responses generated for different simulated users to the same prompt, while the corpus for the across-stage similarity was defined as the responses from the same user across the three stages.

### Semantic Similarity Analysis

We used a fine-tuned transformer-based model DeBERTa-v3-large-MNLI to assess the semantic similarity between pairs of sentences across-users and across-disease stages[50]. DeBERTa (Decoding-Enhanced BERT with disentangled attention) is a pre-trained LLM designed to capture nuanced contextual relationships[51]. The version used in this analysis was fine-tuned on the Multi-Genre Natural Language Inference (MNLI) dataset, enabling it to classify the relationship between sentence pairs as either entailment (semantic alignment) or contradiction (semantic misalignment), with a probability for each label[39, 50].

To calculate the semantic similarity for answers, we compared two documents (e.g., responses (1) across users one, two, and or three; or (2) across T2D stages prediabetes, diagnosed T2D, or diagnosed T2D with complications, denoted and *D_B_* = {*b*_1_,*b*_2_,…,*b_n_*}. We generate sentence pairs (*a_i_*,*b_j_*) cross documents, treating (*a_i_*) as the premise and (*b_j_*) as the hypothesis. Sentences were tokenized using the DeBERTaV2Tokenizer and encoded with the DeBERTa-v3-large-MNLI model. The model produced logits for entailment and contradiction, and the softmax function was applied to obtain entailment probabilities. These probabilities were constructed into a unidirectional entailment matrix *E_A_*_←*B*_ ∈ *R^m^*^×*n*^, where *E_A_*_←*B*_(*i*,*j*) is the probability that *a_i_* is entailed by *b_j_*. A reverse matrix *E_B_*_←*A*_ ∈ *R^n^*^×*m*^was computed simultaneously. The bi-directional sentence-level entailment matrix was defined as[52]

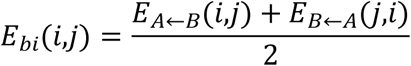

To compare the paragraph-level semantic consistency, we treated the bi-directional sentence-level scores in *E_bi_* as the similarity matrix for alignment. We applied a single-paired alignment method using the Hungarian algorithm (linear sum assignment), which yields the optimal one-to-one pairing that maximizes the total similarity score across all matches[52, 53]. We normalized cumulative similarity scores by the average number of sentences in *D_A_* and *D_B_* to account for differences in text length. For each aligned sentence pair, we used the corresponding *E_bi_*(*i*,*j*) value to classify similarity strength: weak (<0.50), moderate (0.50-0.80), and strong (>0.80). The 0.50 reflects the decision boundary of the two-way model we used, where values larger than 0.50 indicate entailment is more likely than contradict and neutral[50].

We selected 0.80 as a descriptive cutoff to summarize sentence alignment in the strong similarity range. This threshold was not intended to define clinical correctness, it was selected as a pragmatic descriptive reference point informed by previous consistency workflows and prior evaluations showing substantial agreement between entailment estimators and human sentence-level entailment judgments[39, 54, 55]. To reduce reliance on a single threshold, we reported the full distribution of similarity scores in **Figure 1** and **Figure 2**. The summarization of similarity score was reported in **Supplementary table 1 S1** for dietary prompts and **Supplementary table 2 S2** for physical activity prompts. We also summarized common thematic patterns at each similarity band as displayed in **Supplementary table 5 S5**.

### Procedure

We calculated word and sentence similarity for all possible pairwise comparisons across users (i.e., User1 vs. User2, User1 vs. User3, User2 vs. User3) and across Pre-Diabetes, T2D, and T2D with Complications disease stages (i.e., Pre-D vs T2D, Pre-D vs T2D+CKD, T2D vs T2D+DR, T2D+CKD vs T2D+DR, etc.). For stage-specific similarity analysis, the analysis was performed for each user separately and aggregated by mean.

Once all estimates were calculated, we evaluated across-user and across-stage variability using mean difference across prompt and pairwise stage-pair with a user-nested (n = 3) mean-difference bootstrap. Each bootstrap used 10,000 iterations. The results were displayed in **Supplementary table 3 and 4 (S3, S4).** Bootstrap was conducted under Python (v3.12) using numpy (2.4.2) and pandas (3.0.1).

## Declarations

### Ethics approval

This study did not involve any interaction with human subjects and was deemed non-human subjects research by the Institutional Review Board at Indiana University.

### Consent to participate

This study asked three researchers affiliated with the study to directly query GPT-4o with the same questionnaire. There was no interaction between study researchers and any human subjects.

### Human Ethics and Consent to Participate declarations

Not applicable.

### Clinical trial registration

Clinical trial number not applicable.

### Data availability

The data and code supporting the findings of this study are publicly available at: https://github.com/dvaldez44/GPT_User_Consistency.

### Author contributions

Y.Z. conceived the study, designed the analysis, and drafted the manuscript. X.L. and Q.H. contributed to data collection. K.G. and I.C. assisted in manuscript revision as diabetic expert.

J.C. assisted in the method development. D.V. supervised this study and assisted with manuscript revision. All authors reviewed and approved the final version of the manuscript.

## Acknowledgment

The authors thank all collaborators and data contributors for their valuable support. AI was only used to generate the answers for assessment. AI was not used for manuscript preparation or data analysis.

